# Determinants of Hepatitis B Virus infection Acquisition among Apparently Healthy Pregnant Women Attending RCH Clinics in Ifakara, Tanzania

**DOI:** 10.1101/2025.09.08.25335334

**Authors:** Fauster M Mlilimko, Philbert Balichene Madoshi

**Affiliations:** St. Francis University College of Health and Allied Sciences, Department of Internal Medicine and Clinical Pharmacology, P. O. Box 175, Ifakara, Tanzania; St. Francis University College of Health and Allied Sciences. Department of Public Health, P. O. Box 175, Ifakara, Tanzania

**Keywords:** Ifakara, HBV, SFUCHAS, LMICs, Screening, Pregnant women

## Abstract

**Introduction:** Hepatitis B and C virus (HBV and HCV) infections remain major global public health concerns, contributing significantly to morbidity and mortality. During pregnancy, these infections are associated with adverse outcomes, including gestational diabetes, preterm labor, and low birth weight. Despite the availability of effective vaccines and antiviral therapy, awareness of HBV and HCV status among pregnant women in resource-limited settings remains low. This study aimed to identify risk factors associated with HBV infections among pregnant women attending reproductive and child health (RCH) clinics at a tertiary health facility.

**Methodology and Results:** A cross-sectional study was conducted using simple random sampling to select 385 pregnant women. Data were collected using structured checklists and analyzed with SPSS. The majority of participants (66.2%) were aged 18–29, with most being married (63.9%). Nearly half (48.3%) were self-employed, and 46.2% had attained at least secondary education. Medical and behavioral risk factors were common: 55.3% reported a history of hospital admission, 34.5% had undergone surgery, and 29.9% had received blood transfusions. Additionally, 41.6% had tattoos or body piercings, and 70.7% reported having multiple sexual partners. Other notable findings included a history of sexually transmitted infections (25.7%), intravenous drug use (7.8%), and HIV positivity (14.3%). Despite these risk factors, only 0.52% reported a prior diagnosis of hepatitis C.

**Conclusion:** The findings emphasize the need for routine HBV and HCV screening in antenatal care, particularly for high-risk groups. Strengthening infection prevention, raising awareness, and integrating HBV and HCV testing into maternal health services could reduce transmission. Given the high prenatal care attendance (88.3%), policy changes supporting comprehensive screening and education

## Introduction

Hepatitis B virus (HBV) infection remains a significant global public health concern, affecting an estimated 58 million people worldwide (WHO, 2023). Chronic HBV infection can lead to serious liver complications, including cirrhosis and hepatocellular carcinoma, and poses heightened risks for pregnant women and their unborn children (Pan et al., 2005; Lao et al., 2003). In paediatric populations, vertical transmission is the most common route of infection, with approximately 6% of infants born to HBV-positive mothers acquiring the virus during pregnancy or childbirth (MacLachlan & Cowie, 2015). HBV is a DNA virus transmitted primarily through exposure to infected blood and bodily fluids and can cause both acute and chronic liver disease. Among pregnant women, HBV infection presents considerable risks to maternal health and foetal outcomes. Vertical transmission especially during childbirth is the most common route of infection in high-prevalence regions such as sub-Saharan Africa and Southeast Asia (WHO, 2023; MacLachlan & Cowie, 2015). Chronic HBV infection during pregnancy has been linked to complications such as gestational diabetes, preterm birth, and low birth weight (Lao et al., 2003). Without timely prophylaxis, up to 90% of infants born to HBV-positive mothers may develop chronic infection, particularly when maternal viral loads are high and preventive measures like vaccination and hepatitis B immunoglobulin (HBIG) are not provided (Pan & Zhang, 2005).

The World Health Organization recommends routine HBV screening in antenatal care, followed by appropriate management to prevent mother-to-child transmission (MTCT), a key strategy in reaching the 2030 hepatitis elimination targets (WHO, 2020). Despite the availability of effective prevention tools, many HBV infections remain undiagnosed in resource-limited settings. In high-income countries, common transmission modes include intravenous drug use, haemodialysis, blood transfusions, needle-stick injuries, tattooing, sexual contact, and perinatal transmission. In contrast, in low-resource settings, unsafe medical practices such as the reuse of needles and poor sterilization of instruments significantly contribute to the continued spread of HBV (Pan et al., 2005; WHO, 2023).

Unsafe injection practices are estimated to account for 70% to 95% of HBV infections in low-resource settings, substantially contributing to chronic illness, disability, and mortality (WHO, 2023). Mother-to-child transmission (MTCT) of HBV occurs in approximately 6% of cases; however, the precise mechanisms and timing of this transmission whether in utero, during delivery, or postpartum—are still not fully understood. Studies suggest that the risk of vertical transmission is higher in HBV-infected mothers, especially those with high viral loads, compared to uninfected individuals (Pan et al., 2005; Lao et al., 2003).

The prevalence of HBV among pregnant women varies widely across regions and populations, influenced by socio-demographic, behavioural, and healthcare-related factors (MacLachlan & Cowie, 2015; Ofori-Asenso & Agyeman, 2016). Pregnant women constitute a critical target group for HBV screening due to the risk of vertical transmission, which can lead to chronic infection in the newborn and adverse maternal outcomes. Transmission can occur during pregnancy, childbirth, or potentially through breastfeeding, especially in the absence of timely immune-prophylaxis (WHO, 2020; Pan et al., 2005). Identifying and addressing risk factors among pregnant women is therefore essential for informing public health strategies and improving maternal and neonatal health. In Tanzania, however, there is limited data on the specific risk factors contributing to HBV infection among pregnant women attending Reproductive and Child Health (RCH) clinics, underscoring the need for localized research and targeted interventions.

In Tanzania, the overall prevalence of HBV in the general population is estimated to be below 2% (WHO, 2021); however, there is a paucity of data specifically addressing HBV prevalence and associated risk factors among pregnant women.. This study aimed to identify the risk factors associated with HBV infection among pregnant women attending Reproductive and Child Health (RCH) services at a tertiary healthcare facility. Understanding these risk factors in this vulnerable group is essential for informing targeted public health interventions, particularly in resource-limited settings. Additionally this study will inform evidence-based policy and clinical guidelines, ultimately improving maternal and child health outcomes in Tanzania.

## Methods and materials

### Study area, design and population

The study was conducted at St. Francis Referral Hospital (SFRH), which is located in Ifakara Town Council (ITC), Kilombero District, Morogoro Region, Tanzania. ITC is part of the Kilombero Valley, which comprises of three other administrative councils: Mlimba, Malinyi, and Ulanga. SFRH serves a wide catchment area, offering comprehensive maternal and child health services to both rural and urban populations. This was a health facility-based, descriptive cross-sectional study aimed at investigating the determinants of hepatitis B and C virus (HBV and HCV) infections among apparently healthy pregnant women attending reproductive and child health (RCH) services. The study was conducted for a period of 9 months {February – October 2025].

### Inclusion and exclusion criteria

The inclusion criteria in this study were pregnant women attending Reproductive and Child Health (RCH) clinics at St. Francis Referral Hospital or affiliated facilities in Ifakara, willingness to provide informed consent, being resident of Kilombero valley for at least six months prior to data collection and availability for an interview. The exclusion criteria were critically ill or having medical complications which could hinder participation or communication during data collection. Have known mental illness or cognitive impairments which would affect their ability to consent or respond reliably, and declining to provide informed consent for sample collection.

### Sample size

The sample was calculated using a Cochran formula with assumption of 50% prevalence, 95% confidence level (Z = 1.96) and a margin error of 5% (α = 0.05) at 95% confidence interval. Therefore, the calculated sample size was 384 with an addition of 10% for non-response, the sample size was 422. However at the end of the study only 385 pregnant women were randomly allocated and sampled using the fifth attendee from the ante-natal clinic (ANC) register on a particular day.

### Data collection tools and procedures

In this study primary data were collected from pregnant women attending the RCH clinic at St. Francis Referral Hospital, the data were collected using a pre-tested structured checklist. The checklist was administered through face-to-face by one of the authors who is a trained medical student assisted by a trained nurse. The checklist included (socio-demographic variables such as age, level of education, marital status, occupation, place of domicile), cultural, obstetrical and medical history which included gravidity, number of parity, blood transfusion and surgical procedures, risk factors were tattoos, needle-piercing, substance use and others and knowledge, attitudes and practices on HBV and HCV.

### Quality control

Data quality was ensured by the enumerators whereby the English questions were translated to Swahili then back to English to ensure consistent of the intended meaning. Each question on the checklist was reviewed if the responses were filled before letting the participant leave and departing the study area. Any checklists that were not properly filled out were returned back to participants for correction. All completed checklist were entered into a secure, password protected database and cleaned before the analysis was conducted. In this study the confidentiality was strictly maintained by removing all personal identifies.

### Data analysis procedures

The data were analysed using SPSS version 26 (IBM Corp. Armonk, NY, USA), the descriptive statistics: frequencies and percentages were used to summarise categorical variables. Chi – square test was used to assess the relationship and distribution of the outcome on independent variables. The results are presented with a p – value < 0.05 was considered as statistically significant.

### Ethical consideration

Ethical clearance for the study was obtained from the Research and Ethical Committee of St Francis University College of Health and Allied Sciences (SFUCHAS). Written signed consent was received from every participant before recruitment into the study. Participants were informed on their rights to participate or withdraw from the study at any time they feel to. Participants were identified by unique numbers and not names to strictly maintain confidentiality.

## Results

The study recruited 385 pregnant women attending the Reproductive and Child Health (RCH) clinic at St. Francis Referral Hospital in Ifakara. Data were collected using a structured checklist, achieving a 100% response rate. After data cleaning, all entries were deemed valid for analysis. The checklist captured demographic, medical, cultural, and obstetrical risk factors associated with hepatitis B and C virus (HBV and HCV) acquisition. Demographically, the majority of participants were aged 18–24 years (35.6%), In terms of marital status, 63.9% were married. With regard to education, 46.2% had completed secondary education.. Economic status showed that 48.3% of participants had informal employment. (**Figure 1**). In the addition the statistical inferences present that age-group (p = 0.020) showed significant differences. Whereas education level (p = 0.08), marital status (p = 0.754) and occupational status (p = 0.667) were not different statistically at 95% confidence level.

**Figure 1.**
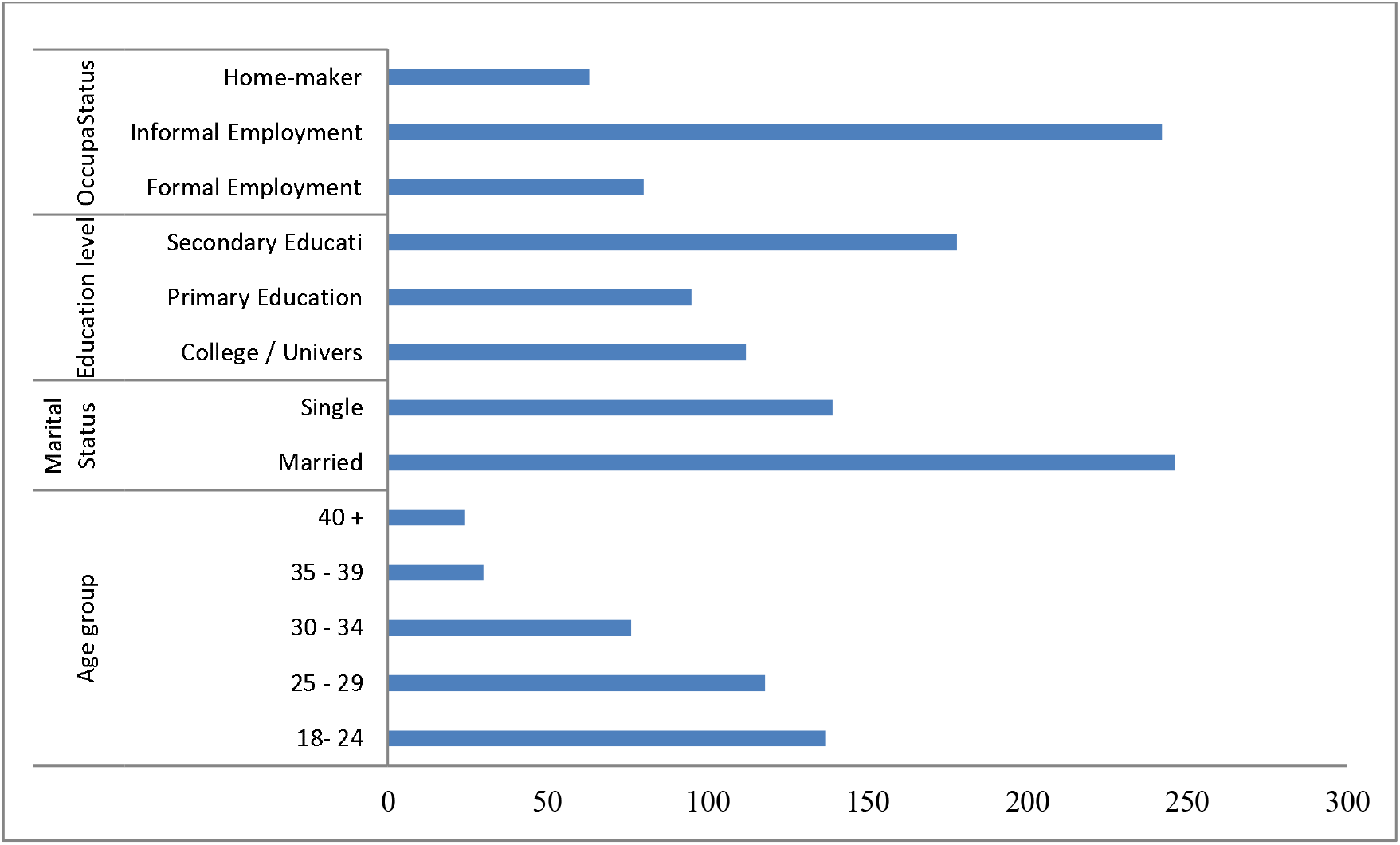
Demographic presentation of the participants.

### Medical associated risk factors

The study also assessed participants’ medical histories for risk factors associated with HBV and HCV infections. A total of 29.9% (115 women) reported having received a blood transfusion, while 34.5% had undergone surgical procedures both of which are known risk factors for HBV transmission. Additionally, 55.3% had been hospitalized at least once, often due to medical conditions that could increase exposure risk. Notably, 3% of participants reported a previous diagnosis of HBV infection, and 35.3% had a family history of liver disease. Furthermore, 14.3% of respondents were living with HIV, 6.2% had a history of hemodialysis, and 7.8% reported intravenous drug use. A history of sexually transmitted infections (STIs) was reported by 25.7% (99 women) (**Table 1**).

**Table 1.** Medical risk factors associated with HBV and HCV acquisition among participants

| Variables | Description | Distribution (N, %) | *p* - value |
| --- | --- | --- | --- |
| Previous History of blood transfusion | Employed Formal | 80 (20.8%) | *0.020* |
|  | Employed Informal | 56 (14.5%) |  |
|  | Self employed | 186 (48.3%) |  |
|  | Unemployed | 63 (16.4%) |  |
| History of surgical operations | No | 270 (70.1%) | *0.444* |
|  | Yes | 115 (29.9%) |  |
| History of hospitalisation | No | 252 (65.5%) | *0.705* |
|  | Yes | 133 (34.5%) |  |
| History of hepatitis B and C virus Infections | No | 172 (44.7%) | *0.508* |
|  | Yes | 213 (55.3%) |  |
| History of Sexual Transmitted Infections | No | 113 (29.4%) | *0.048* |
|  | Yes | 272 (70.6%) |  |
| History of Family liver disease | No | 345 (89.6%) | *0.834* |
|  | Yes | 40 (10.4%) |  |
| HIV status | No | 249 (64.7%) | *0.558* |
|  | Yes | 136 (35.3%) |  |
| History of Haemodialysis | Negative | 330 (85.7%) | *0.044* |
|  | Positive | 55 (14.3%) |  |
| History if IV Drug Administration | No | 361 (98.8%) | *0.884* |
|  | Yes | 24 (6.2%) |  |

### Cultural practices leading to HBV infections

The results on the cultural practices associated with the risk of HBV and HCV infections acquisition among pregnant women was assessed. A total of 41.6% of the women reported having a history of tattoos or body piercings practices that may increase the risk of blood-borne infections. Additionally, 71.2% of the participants reported having multiple sexual partners, a known risk factor for HBV transmission. The use of traditional medicine was also common, with 64.4% of participants reporting its use. This practice was more prevalent among women aged 30–34 (19.7%) and 35–39 (7.8%), who often perceived traditional remedies as the first line of treatment for health complications during pregnancy.

### Obstetrical behaviour leading to HBV infection acquisition

This category assessed obstetric and gynecological factors such as parity, history of miscarriage, preterm delivery, antenatal clinic (ANC) attendance, and gestational diabetes. The distribution of parity showed that 22.6% of participants were in their second pregnancy; while a higher proportion (28.6%) were in their third. A history of miscarriage was reported by 16.9% of the women, and 37.1% had experienced at least one preterm birth. Most participants (88.3%) reported regular attendance at ANC services. Additionally, 4.7% had a history of gestational diabetes (**Table 2**).

**Table 2.** Obstetrical behaviour leading to HBV infection acquisition

| **Variables** | **Description** | **Frequency** | ***p - value*** |
| --- | --- | --- | --- |
| Parity status | No | 137 (35.6%) | *p = 0.770* |
|  | Yes | 248 (64.4%) |  |
| History of miscarriage | First pregnancy | 97 (25.2%) | *p = 0.608* |
|  | Second pregnancy | 87 (22.6%) |  |
|  | Third pregnancy | 110 (28.6%) |  |
|  | Four or more | 91 (23.6%) |  |
| History of preterm delivery | No | 320 (83.1%) | *p = 0.986* |
|  | Yes | 65 (16.9%) |  |
| Ante natal clinic attendance | No | 242 (62.9%) | *p = 0.571* |
|  | Yes | 143 (37.1%) |  |
| History of gestation diabetes | No | 45 (11.7%) | *p = 0.014* |
|  | Yes | 340 (88.3%) |  |

## Discussion

This study intended to determine the factors associated with hepatitis B and C virus infections. The study deployed a questionnaire survey in apparently healthy pregnant women attending ante-natal clinic services at one of the referral hospitals in the northeast of Tanzania. The authors hypothesized that if pregnant women are vulnerable to these viral infections, they will also affect their newborns. Furthermore studies combining awareness, knowledge and practices with regards to HBV and HCV are limited in the developing countries. The study included several variables such as socio-demographic, medical, behavioral, and obstetric risk factors influencing HBV and HCV prevalence among pregnant women. These insights provide critical implications for maternal healthcare interventions, HBV and HCV screening programs, and infection prevention strategies.

Socio-demographic factors such as age, gender, educational level, and occupation are known to influence the acquisition of HBV and HCV infections across different populations. In the present study, younger adults—particularly those aged 18–24 years—comprised the majority of participants and showed statistically significant associations with infection. These findings align with studies from Ethiopia and Nigeria, where younger individuals were more likely to engage in high-risk behaviors such as unsafe sexual practices and traditional tattooing (Belyhun et al., 2011; Forbi et al., 2010). Similarly, Ofori-Asenso and Agyeman (2016) noted that individuals in this age group often have greater behavioral exposure risks. Conversely, Ullah et al. (2013) highlighted that younger women may also be vulnerable to unsafe medical practices during childbirth, increasing their risk of HBV/HCV. In contrast, this study did not find a significant association between educational level, occupation, or marital status and infection, which differs from previous studies in Tanzania and Egypt where low educational attainment was linked to increased infection risk due to limited health literacy (Mwakapotera et al., 2017; Talaat et al., 2010). These comparisons emphasize the role of socio-demographic disparities in shaping viral hepatitis exposure and underscore the importance of localized public health interventions.

Medical-associated risk factors such as history of blood transfusion, surgical procedures, dental treatments, and lack of screening during antenatal care are significant contributors to hepatitis B and C virus (HBV and HCV) infections among pregnant women. Our study could not establish the statistical association of hospitalization history, history of surgical operation; family history of liver diseases or intravenous drug administration. Our study contravenes other studies which s have shown that inadequate knowledge and poor preventive practices among pregnant women amplify these risks. Tadesse et al. (2020) in Ethiopia reported that pregnant women unaware of HBV transmission through medical procedures were more likely to engage in high-risk practices. Odinaka et al., (2013) in Nigeria highlighted that poor awareness and attitudes toward infection control during antenatal visits increased exposure to unsafe medical interventions, such as unscreened blood transfusions and shared medical instruments (). While Mwakapotera et al. (2017) in Tanzania reported a small proportion of pregnant women that receive HBV screening during routine antenatal care, despite high-risk histories, suggesting significant gaps in both practice and policy. These findings underscore the need for health education and strengthened antenatal protocols to address medical-related risk factors and reduce HBV/HCV transmission.

Cultural practices play a significant role in the transmission dynamics of HBV and HCV. In this study, cultural factors such as the use of traditional medicines, tattooing or body piercing, and having multiple sexual partners were assessed. Although these variables were not statistically associated with HBV or HCV infection among pregnant women, their epidemiological relevance should not be underestimated. In remote or underserved communities, limited health literacy and adherence to traditional practices may still pose a significant risk. For instance, Okonkwo et al. (2019) in Nigeria reported increased HCV seropositivity among women with traditional tattoos or piercings. Similarly, Ngemera et al. (2021) in Tanzania linked cultural initiation rites to higher HBV prevalence, especially in rural areas with poor antenatal coverage and informal education. Tugizov et al. (2014) and Oti et al. (2016) also noted high HBV and HCV prevalence linked to exposure through non-sterile cultural practices. Moreover, studies by Adoga et al. (2010) and Adekanle et al. (2015) revealed low hepatitis awareness among pregnant women using traditional herbal medicine. These findings emphasize the need for targeted health education campaigns and culturally sensitive interventions during antenatal care to reduce both vertical and horizontal transmission of viral hepatitis.

These are significant contributors to the acquisition and transmission of hepatitis B (HBV) and hepatitis C (HCV) among pregnant women, particularly in low-resource settings. In our study, factors such as parity, history of miscarriage, preterm delivery, and antenatal care (ANC) attendance were not statistically associated with HBV or HCV infections. However, a history of gestational diabetes was significantly associated with increased infection risk, potentially due to frequent medical interventions and compromised immunity. These findings diverge from those of Damtie et al. (2021) in Ethiopia, who found that surgical procedures and blood transfusions were strongly linked to HBV infection due to inadequate infection control practices [Damtie et al. 2021]. Similarly, Ali et al. (2017) in Pakistan reported that poor awareness of sterile procedures during gynaecological care contributed to elevated HCV rates [Ali et al. 2017]. Mwakapeje et al. (2020) in Tanzania highlighted that misconceptions and low knowledge about HBV among pregnant women negatively influenced their health-seeking behavior [Mwakapeje et al. 2020]. Moreover, research by Umar et al. (2022) indicated that repeated pelvic procedures without adherence to standard precautions increased HBV risk in obstetric settings (Umar et al. 2022). These findings underscore the need for integrating HBV and HCV education into ANC services, enforcing universal precautions among healthcare workers, and enhancing infection prevention during obstetric and gynaecological care.

Furthermore, parity has been identified as a potential risk factor for hepatitis B virus (HBV) and hepatitis C virus (HCV) infections among pregnant women, particularly in low-resource settings where healthcare services may be inconsistent and awareness of transmission risks is limited. Multiparous women are often more exposed to obstetric interventions such as deliveries, cesarean sections, and blood transfusions, which increase their cumulative risk of HBV and HCV exposure. A study in Nigeria by Adekanle et al. (2015) revealed a significantly higher HBV prevalence among women with higher parity, likely due to repeated contact with healthcare settings where infection control practices may be suboptimal. Similarly, research conducted in Sudan by Gasim et al. (2017) demonstrated an association between high parity and increased risk of HCV infection, suggesting that cumulative exposure to invasive procedures may contribute to transmission. Moreover, limited knowledge and misconceptions regarding infection risks during childbirth among multiparous women were highlighted in a study from Tanzania by Msuya et al. (2020), where a majority of participants lacked awareness of hepatitis transmission modes. These findings underscore the need for enhanced health education targeting multiparous women during antenatal visits to improve attitudes and promote safe maternal practices.

## Conclusion

This study identified key determinants associated with Hepatitis B virus (HBV) infection among apparently healthy pregnant women attending RCH clinics in Ifakara, Tanzania. Although most socio-demographic and obstetric factors were not statistically significant, medical risk factors such as a history of blood transfusion, gestational diabetes and haemodialysis showed strong associations with HBV infection. These findings underscore the importance of routine HBV screening during antenatal care, enhanced public health education, and strengthened infection prevention measures. Targeted interventions are essential to reduce HBV transmission and improve maternal and neonatal health outcomes in resource-limited settings like Ifakara.

## Data Availability

All data produced in the present study are available upon reasonable request to the authors

## Acknowledgement

The authors acknowledge the administration of St. Francis Referral Hospital for the facilitating data collection, the Institutional Review Board of St. Francis University College of Health and Allied Sciences for the support and the nurses at the RCH clinics.

## Data availability statement

The data supporting the findings of this study are available from the corresponding author upon reasonable request.

## Ethical clearance

This study was approved by the Institutional Review Board of St. Francis University College of Health and Allied Sciences ((IRB-SFUCHAS-2023-001), Furthermore, this study was approved by the research unit of St. Francis Referral hospital and each participant signed an informed consent form before being interviewed.

## Authors’ contribution

1. Philbert Balichene Madoshi: Study conceptualization, data analysis, final document review and approval.
2. Hilda G. Swai: Questionnaire enumeration, data collection and entry, manuscript draft and initial data analysis
3. Jovin Tibenderana: Final data analysis, document review and final document review

## Notes

### Competing Interest Statement

The authors have declared no competing interest.

### Funding Statement

This study did not receive any funding

### Author Declarations

St. Francis University College of Health and Allied Sciences, Institutional Review Board. Approved study

